# The impact of the COVID-19 pandemic on health related quality of life in head and neck cancer survivors: an observational cohort study

**DOI:** 10.1101/2023.01.03.23284145

**Authors:** B.I. Lissenberg-Witte, F. Jansen, R.J. Baatenburg de Jong, F. Lamers, C.R. Leemans, S.F. Oosting, R.P. Takes, I.M. Verdonck-de Leeuw

## Abstract

**Background:** Physical, psychological, and social aspects of health-related quality of life (HRQOL) among head and neck cancer (HNC) survivors may be more affected during the COVID-19 pandemic than before the pandemic. However, the impact is not yet understood well.

**Methods:** Prospectively collected data from the NETherlands QUality of life and BIomedical Cohort study in HNC were used. All patients were diagnosed and treated before the COVID-19 pandemic. Patient reported outcome measures (PROMs) collected 24 and 36 months after treatment (M24 and M36) were compared between survivors who completed both assessments before the COVID-19 pandemic and those who completed M24 before but M36 during the pandemic. Personal, clinical, physical, psychological, social, and lifestyle characteristics of the survivors assessed at baseline or M24 were investigated as potential effect modifiers.

**Results:** In total, 318 HNC survivors were included, of which 199 completed both M24 and M36 before the COVID-19 pandemic and 119 completed M24 before but M36 during the pandemic. Changes in HRQOL between 24 and 36 months follow-up did not differ between the two groups for any of the PROMs. However, in some subgroups of HNC survivors the COVID-19 pandemic negatively affected the course of HRQOL for several PROMs while it positively affected the course of HRQOL for other PROMs.

**Conclusions:** The COVID-19 pandemic did not affect HRQOL in HNC survivors in general, but some subgroups were affected in a positive and others in a negative way.

**Funding:** This work was supported by the Dutch Cancer Society [grant number VU 2013–5930] and the Dutch Cancer Society, Alpe Young Investigator Grant [grant number 12820].

## Introduction

Head and neck cancer (HNC) and its treatment negatively affect the physical, psychological, and social aspects of health-related quality of life (HRQOL) not only before and during treatment but also in long-term survivors [1-6]. It may be that HRQOL of HNC survivors is even more affected in the setting of the COVID-19 pandemic which had an enormous health care, societal and economic impact, but this is not yet understood very well. Previous studies among cancer patients reported a negative impact of the COVID-19 pandemic on HRQOL related to physical [7, 8], psychological [7, 9-12], and social functioning [7, 10, 12]. Other studies found no effect of the COVID-19 pandemic on HRQOL [13, 14] or showed an improvement in physical, role, and social functioning [12] and less loneliness [10]. Several studies investigated possible effects of the COVID-19 pandemic on HRQOL among HNC patients specifically [15-19]. Gallo et al. reported a negative effect on physical, role and emotional functioning and no effect on cognitive and social functioning, global quality of life, or cancer related symptoms [15]. Hamilton et al. found a negative impact on cancer related symptoms [16]. Kirtane et al. conducted a qualitative study and found a negative impact of the COVID-19 pandemic on psychological distress and social isolation [17]. In contrast, Rodrigues-Oliveira et al. found no differences on symptoms of anxiety and depression between HNC patients during radiotherapy before or after the COVID-19 pandemic [18]. Büntzel et al. reported a negative effect on physical and psychological function, and isolation, and a positive effect on relations with family and nature [19].

An explanation for these different and sometimes contradictory findings on the impact of the COVID-19 pandemic among cancer patients might be that most investigators used cross-sectional study designs, and compared the results to reference values or historical cohorts [9-11, 13-19]. Only few studies investigated HRQOL longitudinally, in which part of the measurements were carried out before the COVID-19 pandemic and others during the pandemic [8, 12] or in which cancer patients were prospectively followed during the COVID-19 pandemic [7]. Another explanation for the various previous findings might be that the COVID-19 pandemic affected some patients more than others. Factors that might moderate the impact of the COVID-19 pandemic are age, sex, educational level, marital status, household composition, type and stage of cancer, treatment intent, physical health, job security, and pre-existing psychological problems [7, 9, 14]. Thus far, studies longitudinally investigating the effect of the COVID-19 pandemic on HRQOL among HNC survivors are lacking. The aim of this study is to prospectively investigate the impact of the COVID-19 pandemic on HRQOL among HNC survivors. The following two research questions are addressed: 1) Did the COVID-19 pandemic change the course of HRQOL over time (disease and tumor specific HRQOL, physical activity, symptoms of distress, anxiety, and depression, fear of cancer recurrence, and loneliness), 2) Can subgroups of HNC patients (in terms of personal, clinical, psychological, physical, social, lifestyle, and disease-related factors) be identified in which the course of HRQOL is positively or negatively affected as a consequence of the COVID-19 pandemic.

## Methods

### Study design and patients

Data and samples were used from the NETherlands QUality of life and BIomedical Cohort study in HNC (NET-QUBIC), a prospective cohort study among 739 HNC patients. Patients were recruited between March 2014 and June 2018. Patients were included before start of treatment (baseline) and data was collected at baseline (T0) and 3, 6, 12, 24, and 36 months (M3, M6, M12, M24 and M36) after end of treatment. Data was derived from an electronic case report form (eCRF) designed for NET-QUBIC, patient reported outcome measures (PROMs) and fieldwork assessments. Newly diagnosed HNC patients were included in NET-QUBIC if they were i) 18 years or older, ii) treated with curative intent for cancer of the oral cavity, oropharynx, hypopharynx, larynx, or unknown primary, and iii) able to write, read, and speak Dutch. Patients were excluded if they i) were unable to understand the questions or test instructions, ii) had severe psychiatric comorbidities (i.e. schizophrenia, Korsakoff’s syndrome, dementia), or iii) were unable to understand informed consent. In the current study, HNC survivors were included if they completed at least one PROM at M36. Furthermore, only the T0, M24 and M36 PROMs were used. PROMs completed before March 14^th^ 2020 were considered ‘before COVID-19’ while PROMs completed on or after March 14^th^ 2020 were considered ‘during COVID-19’. Consent procedures were approved by the Medical Ethical Committee of Amsterdam UMC location VUmc (METc VUmc 2013.301 (A2018.307)-NL45051.029.13)) and all participating hospitals and followed the Dutch Medical Research Involving Human Subjects Act. All included survivors signed informed consent.

Details of the key components of NET-QUBIC were published previously: the study population (including retention, attrition and potential selection bias), eCRF, the outcome assessment protocol, biobanking protocol, data management (collection and storage), and data and sample dissemination procedures [20, 21]. The STROBE guidelines were adhered when reporting the results of this study [22]. As this was a not pre-planned post-hoc analysis of the original NET-QUBIC, no formal sample size calculation for the current study has been performed.

### Outcome measures

Disease specific and tumor specific HRQOL were measured with the European Organisation for Research and Treatment of Cancer (EORTC) Quality of Life Questionnaire (QLQ)-C30 and EORTC-QLQ-HN35, respectively. The EORTC QLQ-C30 is a 30-item questionnaire and consists of one global quality of life scale (QL), five functional scales (physical, role, emotional, cognitive and social functioning), three symptom scales (fatigue, nausea and vomiting, and pain) and six single item symptoms (dyspnoea, insomnia, appetite loss, constipation, diarrhoea and financial difficulties). A sum score (SumSC) is based on the five functional scales, the three symptom scales and five of the six single items (financial toxicity is not included) [23-25]. The EORTC-QLQ-HN35 consists of seven HNC specific symptoms (pain, swallowing, senses, speech, social eating, social contact, and sexuality) and ten single item symptoms (problems with teeth, dry mouth, sticky saliva, cough, opening the mouth wide, weight loss, weight gain, use of nutritional supplements, feeding tubes, and painkillers) [26]. QLQ-C30 and QLQ-HN35 Items were scored on a 4-point Likert scale and scores for each of the subscales range from 0 to 100, where higher scores on QL, SumSC and the functional scales indicate better HRQOL and functioning while higher scores on the symptom scales indicate more symptoms.

Physical activity was measured with the Physical Activity Scale for the Elderly (PASE). The PASE is a 13-item questionnaire measuring duration and frequency of leisure time, household and work-related physical activities [27, 28]. Subscale scores for each of the domains were calculated as well as a total score. Higher scores on the subscales and total score indicate more physical activity. The total activity score was also categorized as very poor, poor, fair, good, very good and excellent.

Distress and symptoms of anxiety and depression were measured using the Hospital Anxiety and Depression Scale (HADS). The HADS is a 14-item questionnaire measuring emotional distress and includes a total scale (HADS-T) and an anxiety (HADS-A) and depression (HADS-D) subscale [29]. Items were scored on a 4-point Likert scale and scores for each of the subscales range from 0 to 21, where higher scores indicate higher extent of distress, depression or anxiety symptoms.

Fear of cancer recurrence was measured with the Cancer Worry Scale (CWS). The CWS is an 8-item questionnaire measuring concerns about developing cancer or developing cancer again and the effect of these concerns on daily life [30]. Items were scored on a 4-point Likert scale, and a total scale score is calculated by summing all items, resulting in a total scale score ranging from 8 to 32. A higher score indicates higher extent of fear of cancer recurrence.

Loneliness was measured with the De Jong Gierveld Loneliness Scale (De Jong Gierveld). The De Jong Gierveld is an 11-item questionnaire measuring emotional and social loneliness [31, 32]. Items were scored on a 3-point Likert scale. Scores for emotional loneliness range from 0 to 6 and scores for social loneliness range from 0 to 5, where higher scores indicate higher loneliness. A total loneliness score was calculated by summing the scores of the two scales. The total loneliness score was also categorized as not lonely (score 0 to 2), moderate (score 3 to 8), severe (score 9 or 10) and very severe (score 11) [33].

### Influencing factors

Data on personal, clinical, physical, psychological, social, and lifestyle characteristics were collected from eCRF data, PROMs and fieldwork assessments.

Personal factors (assessed at baseline) included age, sex, educational level (low, middle or high), living status (alone or cohabiting), marital status (married or not married), and personality. Personality was assessed by the extraversion subscale of the 60-item NEO Five Factor Inventory (NEO-FFI) questionnaire [34]. Items are scored on a 5-point Likert scale. The extraversion subscale consists of 12 item and ranges from 12 to 60, where a higher score indicates a higher level of extraversion.

Clinical factors included tumor location (oral cavity, oropharynx, hypopharynx, larynx or unknown primary), tumor stage (I/II or III/IV), treatment modality (single or multimodality treatment), World Health Organization (WHO) performance status (0, able to carry out all normal activity without restriction or ≥1, restricted in normal activities), comorbidity and cancer progression at 24 months follow-up. Comorbidity was assessed by the 27-item Adult Comorbidity Evaluation-27 (ACE-27) Index, which categorizes comorbidity as none, mild, moderate and severe [35]. Cancer progression status between end of treatment and M24 was categorized as residual disease, recurrence and/or second primary tumor at M24 or none of those at M24.

Physical impairments in instrumental activities in daily life was assessed by the Instrumental Activities of Daily Living (IADL) questionnaire [36].

Psychological characteristics included the presence of a major depressive disorder in the past year at M24, and presence of a lifetime major depressive disorder at M24. Presence of a major depressive disorder in the past year was assessed with the Composite International Diagnostic Interview (CIDI), which is based on DSM-IV criteria [37]. The CIDI was assessed yearly during the fieldwork assessment, i.e. at T0, M12 and 24. At T0, presence of a lifetime major depressive disorder was also assessed. A lifetime major depressive disorder was scored as present in case it was already present at baseline or when a major depressive disorder in the past year was diagnosed during the CIDI at M12 and/or M24.

Having paid work (yes or no) was assessed by the iMTA productivity cost questionnaire at baseline and at M24.

Lifestyle-related factors included excessive alcohol consumption (categorized as no or yes (at least 14 (women) or 21 (men) glasses of alcohol per week)), smoking behavior (categorized as current smoker or never smoker and former smoker), and body mass index (BMI) at 24 months.

### Statistical analyses

Data is described by number and frequency in case of categorical variables and by mean and standard deviation (SD) in case of continuous variables. Differences in baseline characteristics between survivors who completed M36 before COVID-19 and survivors who completed M36 during COVID-19 as well as between survivors included in the current study and the other NET-QUBIC participants were analyzed by the chi-square test or the independent sample t-test, depending on the distribution of the variable. Difference in change from M24 to M436 of continuous PROMs between the groups were analyzed by linear mixed effects models, with fixed effect for group, follow-up measurement (M24 or M36) and their two-way interaction and a random intercept for subject. Differences in change of dichotomous or categorical PROMs were analyzed by generalized estimating equations (GEE), with a logit-link function (dichotomous PROMs) or cumulative logit-link function (categorical PROMs). Reported effect sizes, with corresponding 95% confidence intervals (CI) for the change between M24 and M36 include the difference between groups in change between M24 and M36 based on the estimated marginal means for continuous PROMs and odds ratios (OR) for dichotomous and categorical outcomes.

The modifying effect of potential influencing factors for the change between M24 and M36 was also analyzed using linear mixed effects model, including fixed effects for group, measurement and the potential effect modifier, all two-way interactions and the three-way interaction as well as a random intercept for subject. In case of multiple effect modifiers for one PROM, analyses were repeated after stratification for the factor with the lowest p-value (below 0.05) for the three-way interaction. Continuous effect modifiers were stratified by median split. Stratification was only done in case each stratum and each group (completely before COVID or partly during COVID) contained at least 20 survivors. All analyses were performed in SPSS version 28 (IBM Corp., Armonk, NY, USA). The two-sided significance level was set at 0.01, to account for the large number of PROMs analyzed.

## Results

### Study population

Of the 739 participants included in NET-QUBIC, 487 were known to be alive and included in NET-QUBIC at M24 of which 345 survivors completed at least one PROM questionnaire at the M36 assessment, 199 before the COVID-19 pandemic and 146 during the pandemic. Twenty-seven of these 146 survivors also filled in the questionnaires at M24 during the pandemic, and were excluded form analyses resulting in 119 survivors in the group with the M36 assessment during the pandemic (Figure 1). Baseline characteristics, stratified by group, are displayed in Table 1. There were no significant difference in characteristics between the groups. HNC survivors included in the current study (n=318) had lower comorbidity and less often a lifetime major depressive disorder at M24 compared to HNC survivors who were not included (n=142) in the current study (Supplementary Table 1).

**Table 1.** Patients characteristics at baseline unless specified otherwise. Data is described as number and percentage or as mean (standard deviation).

|  | M36 assessment |  |  |  | p-value |
| --- | --- | --- | --- | --- | --- |
|  | before COVID-19<br>(N=199) |  | during COVID-19<br>(N=119) |  |  |
| Age (years) | 63.5 (9.6) |  | 63.6 (9.9) |  | 0.89 |
| Gender |  |  |  |  | 0.091 |
| Men | 140 | 70.4% | 94 | 79.0% |  |
| Women | 59 | 29.6% | 25 | 21.0% |  |
| Education level |  |  |  |  | 0.33 |
| Low | 82 | 43.4% | 41 | 35.3% |  |
| Middle | 50 | 26.5% | 32 | 27.6% |  |
| High | 57 | 30.2% | 43 | 37.1% |  |
| Missing | 10 |  | 3 |  |  |
| Living arrangement |  |  |  |  | 0.044 |
| Living together | 157 | 83.1% | 86 | 73.5% |  |
| Living alone | 32 | 16.9% | 31 | 26.5% |  |
| Missing | 10 |  | 2 |  |  |
| Marital status |  |  |  |  | 0.33 |
| Not married | 62 | 32.8% | 45 | 38.5% |  |
| Married | 127 | 67.2% | 72 | 61.5% |  |
| Missing | 10 |  | 2 |  |  |
| NEO-FFI extraversion<sup>a</sup> | 41.2 (6.1) |  | 40.3 (6.5) |  | 0.23 |
| Tumor location |  |  |  |  | 0.69 |
| Oral cavity | 50 | 25.1% | 30 | 25.2% |  |
| Oropharynx | 72 | 36.2% | 43 | 36.1% |  |
| Hypopharynx | 14 | 7.0% | 4 | 3.4% |  |
| Larynx | 57 | 28.6% | 37 | 31.1% |  |
| Unknown primary | 6 | 3.0% | 5 | 4.2% |  |
| Tumor stage |  |  |  |  | 0.19 |
| Stage 0(Cis)/I<sup>*</sup> | 52 | 26.1% | 32 | 26.9% |  |
| Stage II | 35 | 17.6% | 24 | 20.2% |  |
| Stage III | 38 | 19.1% | 12 | 10.1% |  |
| Stage IV | 74 | 37.2% | 51 | 42.9% |  |
| Treatment modality (dichotomized) |  |  |  |  | 0.56 |
| Single modality | 114 | 57.3% | 64 | 53.8% |  |
| Multimodality | 85 | 42.7% | 55 | 46.2% |  |
| WHO performance status |  |  |  |  | 0.59 |
| 0 | 152 | 76.4% | 94 | 79.0% |  |
| >0 | 47 | 23.6% | 25 | 21.0% |  |
| Comorbidity |  |  |  |  | 0.55 |
| None | 68 | 34.7% | 48 | 40.7% |  |
| Mild | 76 | 38.8% | 46 | 39.0% |  |
| Moderate | 36 | 18.4% | 18 | 15.3% |  |
| Severe | 16 | 8.2% | 6 | 5.1% |  |
| Missing | 3 |  | 1 |  |  |
| Cancer progression status at M24 |  |  |  |  | 0.98 |
| Disease free | 165 | 82.9% | 98 | 83.1% |  |
| Residual, recurrence and/or SP | 34 | 17.1% | 20 | 16.9% |  |
| Residual | 5 | 2.5% | 3 | 2.5% |  |
| Recurrence | 18 | 9.0% | 12 | 10.2% |  |
| Second primary | 12 | 6.0% | 7 | 5.9% |  |
| Missing |  |  | 1 |  |  |
| Type tumor recurrence |  |  |  |  |  |
| Local recurrence (< 2 cm and <3 year) | 6 | 33.3% | 3 | 25.0% |  |
| Regional recurrence | 3 | 16.7% | 3 | 25.0% |  |
| Distant metastasis | 3 | 16.7% | 1 | 8.3% |  |
| Delayed lymph node metastasis | 6 | 33.3% | 5 | 41.7% |  |
| Treatment intent (in case of residual, recurrence and/or SP) at M24 |  |  |  |  |  |
| Curative intent | 30 | 15.1% | 19 | 16.1% |  |
| Palliative intent | 4 | 2.0% | 1 | 0.8% |  |
| IADL at M24<sup>b</sup> | 7.3 (1.0) |  | 7.2 (1.0) |  | 0.33 |
| Major depressive disorder past year at M24 |  |  |  |  | 0.54 |
| No | 172 | 95.6% | 109 | 97.3% |  |
| Yes | 8 | 4.4% | 3 | 2.7% |  |
| Missing | 19 |  | 7 |  |  |
| Lifetime major depressive disorder at M24 |  |  |  |  | 0.24 |
| No | 124 | 82.1% | 69 | 75.8% |  |
| Yes | 27 | 17.9% | 22 | 24.2% |  |
| Missing | 48 |  | 28 |  |  |
| Paid work at M24 |  |  |  |  | 0.94 |
| No | 134 | 70.5% | 83 | 70.9% |  |
| Yes | 56 | 29.5% | 34 | 29.1% |  |
| Missing | 9 |  | 2 |  |  |
| Excessive alcohol consumption at M24 |  |  |  |  | 0.38 |
| No | 163 | 88.1% | 95 | 84.1% |  |
| Yes | 22 | 11.9% | 18 | 15.9% |  |
| Missing | 14 |  | 6 |  |  |
| Smoking behavior at M24 |  |  |  |  | 0.79 |
| Not a current smoker | 162 | 87.1% | 97 | 88.2% |  |
| Current smoker | 24 | 12.9% | 13 | 11.8% |  |
| Missing | 13 |  | 9 |  |  |
| BMI (kg/m2) at M24<sup>b</sup> | 25.8 (4.2) |  | 26.2 (4.1) |  | 0.42 |

\*One patient had a cTNM stage of 0, however, pTNM was stage 2.
<sup>a</sup>missing for n=8 before COVID and n=7 during COVID
<sup>b</sup>missing for n=19 before COVID and n=7 during COVID
SP=second primary; NEO-FFI= NEO Five Factor Inventory; IADL= deze Instrumental Activities of Daily Living; BMI=body mass index

**Figure 1.**
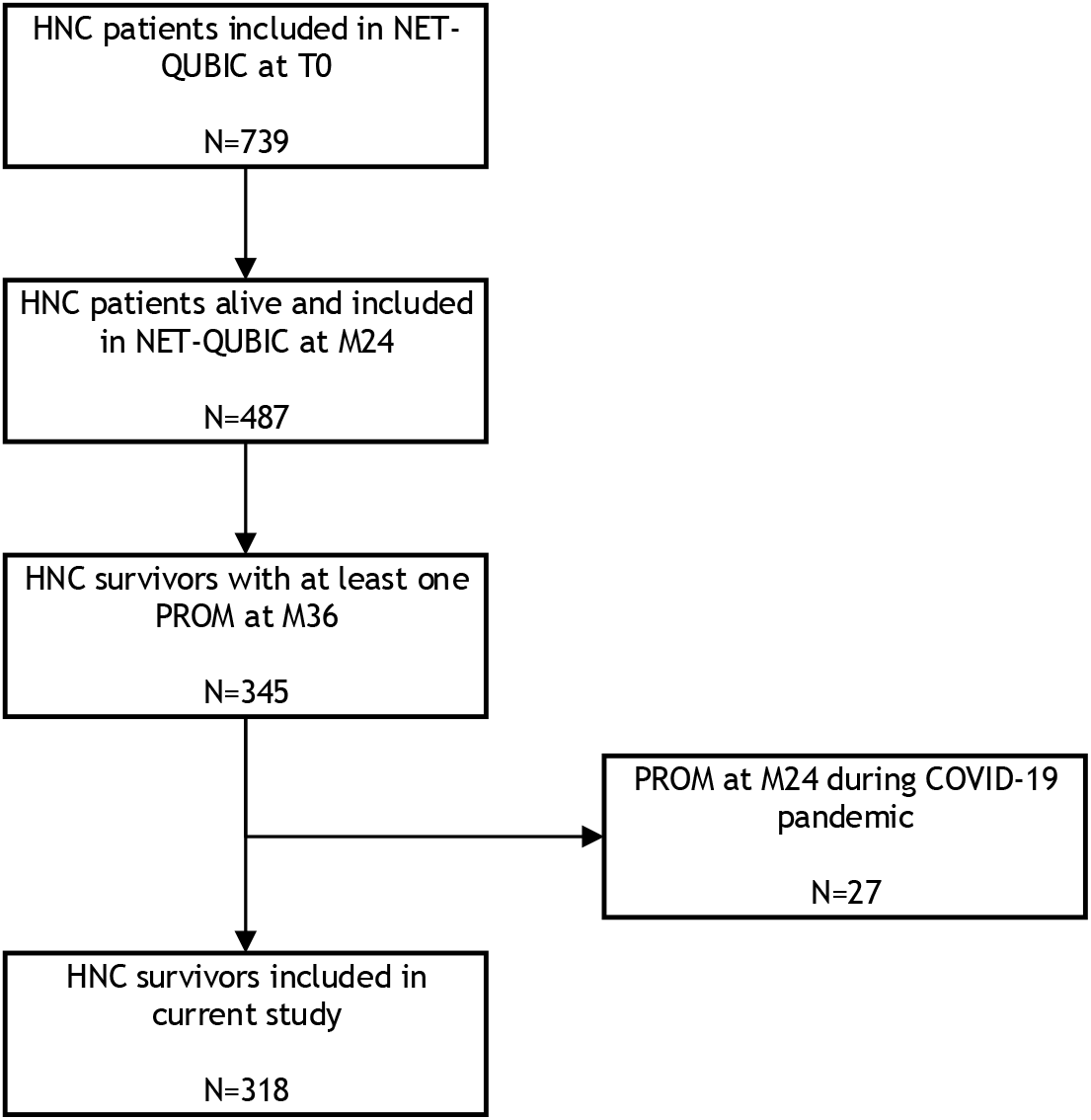
Flow diagram of NET-QUBIC patients included in the current study. HNC=head and neck cancer; M24=24 months follow-up assessment; M36=36 months follow-up assessment; PROM=patient
reported outcome measure; T0=baseline assessment

### The course of HRQOL in relation to the COVID-19 pandemic

There were no statistically significant differences in the change of PROMs between 24 and 36 months follow-up between survivors who completed all PROMs before COVID-19 and those who completed the M36 assessment during COVID-19 (Table 2 and Supplemental Tables 2 and 3).

**Table 2.**
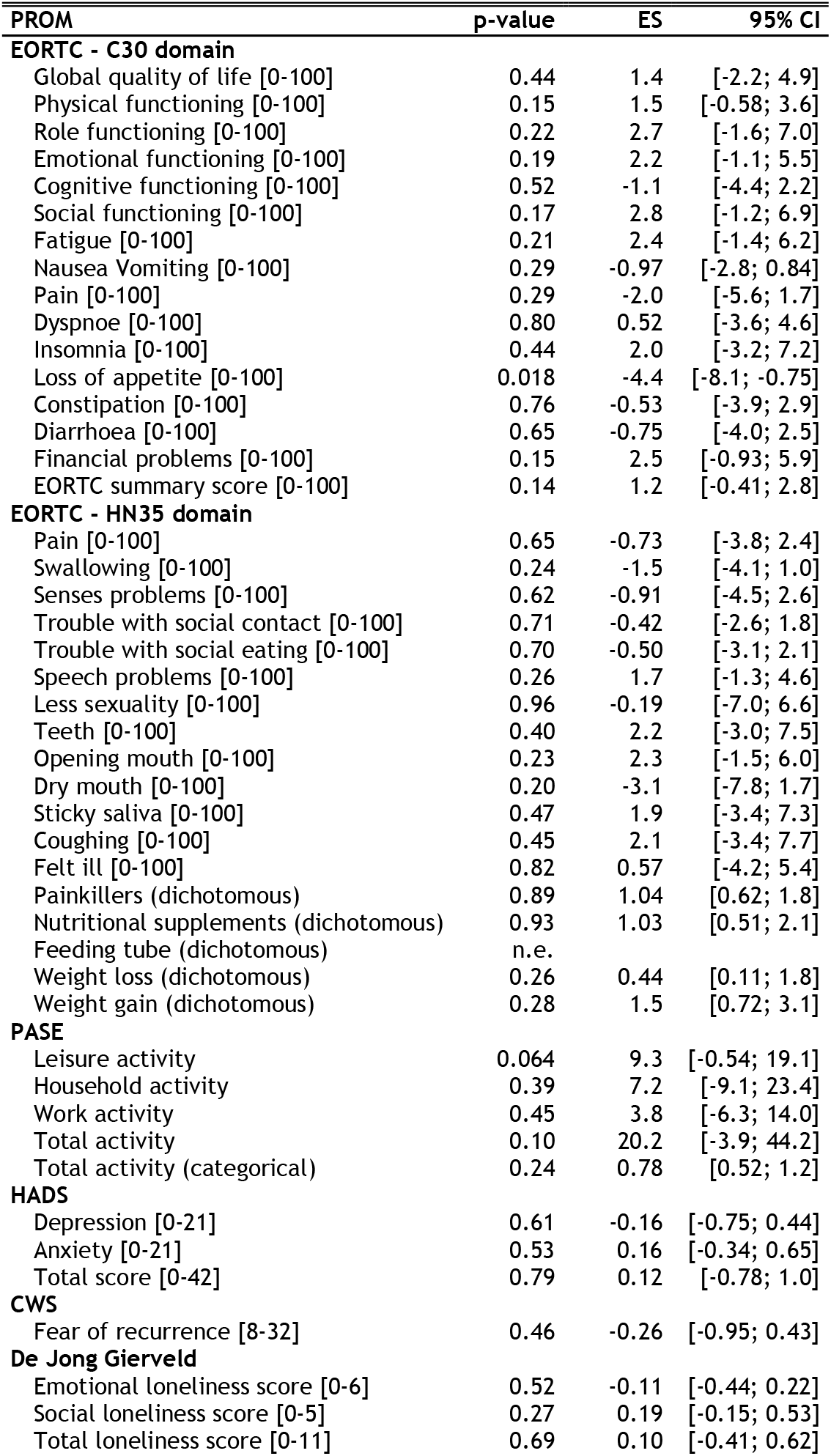
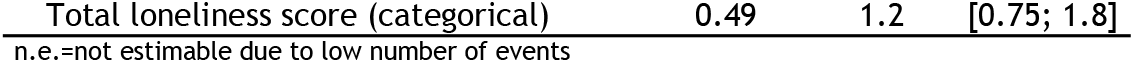
Estimated effect sizes (estimated differences for continuous PROMs or odds ratios for dichotomous and categorical PROMs) with corresponding 95% confidence intervals and p-values of the two-way interactions time by group, to assess differences between groups in change of PROMs between 24 and 36 months follow-up.

For 31 of the 46 PROMs, one or more factors were identified that showed a different effect of the COVID-19 pandemic on the change of HRQOL between M24 and M36 (p<0.05; Supplemental Table 4). After stratification, the most important factors (with a p-value in at least one of the subgroups <0.01) were: gender, living status at baseline, treatment modality, comorbidity, cancer progression status at M24, presence of a lifetime major depressive disorder at M24, and BMI at M24 (Figure 2; Supplemental Table 5). Females deteriorated in PASE total score during COVID-19 while they improved before COVID-19 (categorical) (OR: 0.25, 95% CI: [0.093; 0.68], p=0.007). Survivors living alone at baseline improved on EORTC-HN35 painkiller use during COVID-19 while they deteriorated before COVID-19 (OR: 3.5, 95% CI [1.5; 8.1], p=0.003). HNC survivors treated by multimodality treatment who completed M36 during COVID-19 showed a deterioration on the EORTC-C30 loss of appetite scale whereas survivors who completed M36 before COVID-19 improved (effect size: −8.5, 95% CI [−13.7; −3.4], p=0.001). Survivors with residual disease, recurrence and/or second primary tumor at M24 who completed M36 during COVID-19 improved on the EORTC-C30 financial problems scale while survivors before COVID-19 deteriorated (effect size: 14.9, 95% CI: [5.4; 24.5], p=0.003). Survivors with moderate and severe comorbidity deteriorated on the EORTC-HN35 swallowing scale during COVID-19 while survivors before COVID-19 improved (effect size: −10.2, 95% CI [−16.5; −3.9], p=0.002). Survivors with a lifetime major depressive disorder present at M24 deteriorated in fear of recurrence during COVID-19 while they improved before COVID-19 (effect size: −3.0, 95% CI: [−5.1; − 0.84], p=0.007). Finally, survivors with a BMI at M24 above the median improved on emotional loneliness during COVID-19 while they deteriorated before COVID-19 (effect size: −0.72, 95% CI: − 1.2; −0.28], p=0.002).

**Figure 2.**
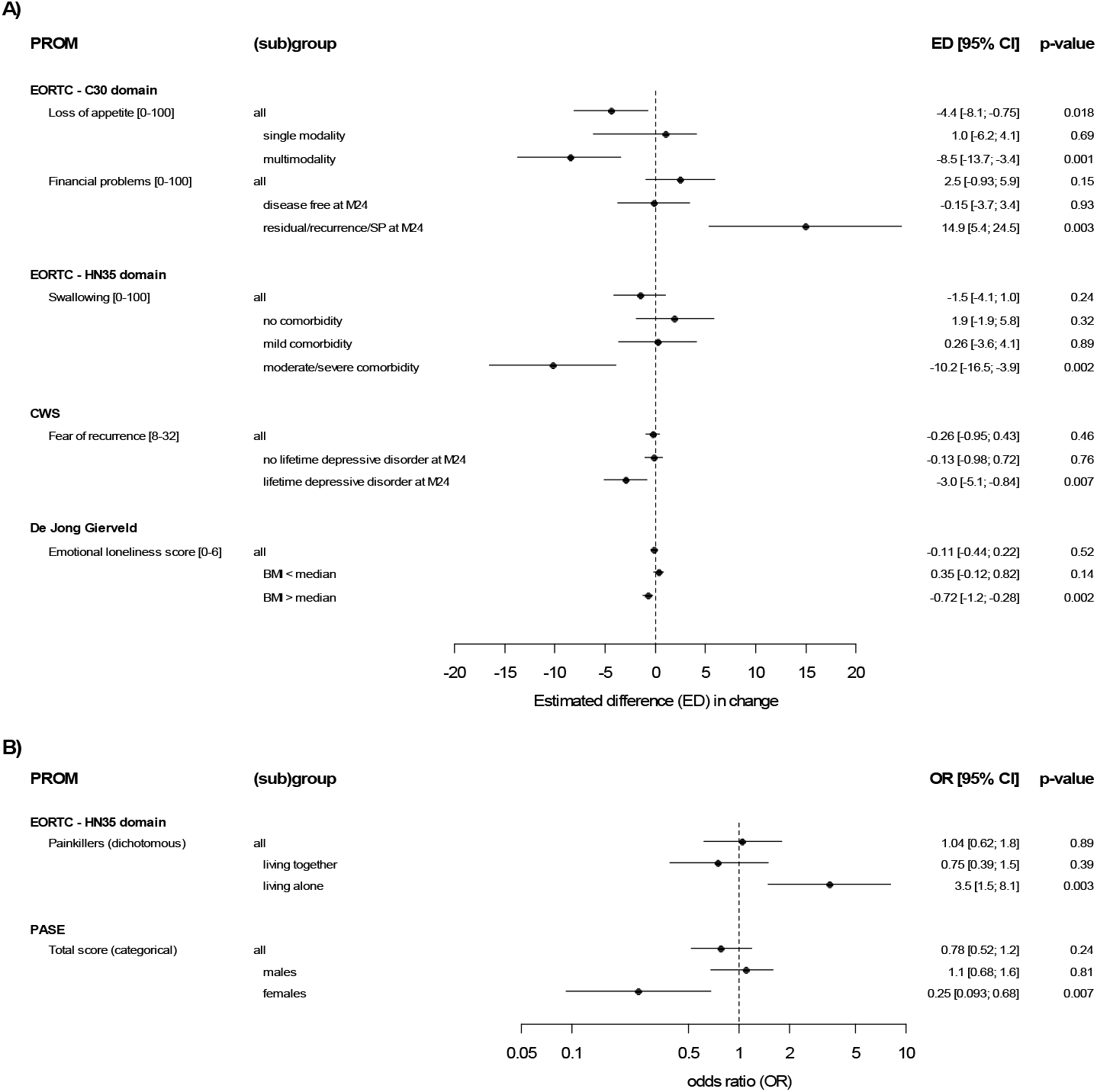
Estimated effect sizes (estimated differences for continuous PROMs or odds ratios for dichotomous and categorical PROMs) with corresponding 95% confidence intervals and p-values for the differences between groups in change of PROMs between 24 and 36 months follow-up, overall and stratified by influencing factor. A) continuous PROMs. B) dichotomous and categorical PROMs.

## Discussion

Over all, the COVID-19 pandemic did not influence the change in HRQOL between 24 and 36 months after treatment in HNC survivors. However, in some subgroups of HNC survivors the COVID-19 pandemic had a negative effect on some PROMs while in other groups there was a positive effect. The COVID-19 pandemic had a negative effect on females (worsening of physical activity), survivors treated with multimodality treatment (worsening of appetite loss), survivors with comorbidity (worsening of swallowing problems), survivors with a history of a major depressive disorder (worsening of fear of recurrence), and survivors with a high BMI (worsening of emotional loneliness). The COVID-19 pandemic had a positive effect on survivors living alone (decrease of painkiller use) and on survivors with disease progression (decrease of financial problems). Previous studies also reported that sex and pre-existing psychological problems moderate the impact of the COVID-19 pandemic [7, 9, 14]. Furthermore, these studies suggested that also age, educational level, marital status, household composition, type and stage of cancer, treatment intent, and physical health might moderate the effect of the COVID-19 pandemic but this was not confirmed by the current study [7, 9, 14].

An explanation for the negative and positive effects of the COVID-19 pandemic among subgroups of HNC survivors might be an altered and unequal access to follow-up or supportive care during the COVID-19 pandemic [7, 8, 17, 38, 39]. Among colorectal cancer survivors in follow-up care during the COVID-19 pandemic, role, emotional and social functioning, fatigue, sleep disturbance, and anxiety was worse in those survivors who had hospital visits canceled, postponed, or changed into digital care, compared with survivors without changes in their cancer care planning [13]. In contrast, a systematic review on studies investigating the effect of a reduction in follow-up frequency during the COVID-19 pandemic among breast cancer patients, showed no adverse effect on HRQOL (nor on survival) but improved cost-effectiveness of follow-up care. Four RCTs that investigated follow-up on-demand versus scheduled follow-up visits found no statistically significant differences in HRQOL [40]. What we learned from the COVID-19 pandemic is that supportive care services were capable to make significant changes in the provision of their care in a short period of time, and that eHealth was often used as integrated part of supportive care. However, it is known that some cancer survivors benefit more from eHealth than others [41]. HNC survivors may also have used peer support but a survey among health care professionals in the UK demonstrated that different types of peer support are available but that referral to peer support is complex and divers [42].

The key strength of this study is the prospective longitudinal research design. A limitation is that all HNC survivors were included and finished their primary treatment for HNC before the COVID-19 pandemic, and the results cannot be generalized to those who were diagnosed and treated during the COVID-19 pandemic. Also, for some variables, data were only available at baseline while their situation may have changed two years later (e.g. marital status). Moreover, we investigated many PROMs, and although we accounted for this by setting the significance level to 0.01, some of our results could have been significant by chance. Finally, we do not know whether participating HNC survivors were diagnosed with COVID-19. Cancer patients in general have higher odds to develop severe COVID-19 and to die of the consequences [43].

In conclusion, the course of HRQOL between 24 and 36 months after treatment in HNC survivors in general was not affected by the COVID-19 pandemic. However, the COVID-19 pandemic did change the course of some HRQOL domains or symptoms over time in some subgroups of HNC survivors. The development of personalized supportive care programs including regular care, eHealth, and peer support, tailored to the needs of the individual survivor may help to overcome disparities among HNC survivors.

## Data Availability

The datasets geneThe dataset generated and analyzed (including a deindentified version of it) during the current study is not publicly available as the collection and integration of large amounts of personal, biological, genetic and diagnostic information precludes open access to the NET-QUBIC research data. Data are available from the corresponding author on reasonable request. On the NET-QUBIC website (www.kubusproject.nl) is described how NET-QUBIC data are made available for the research community. In short, a research proposal has to be submitted to the NET-QUBIC steering committee. The researcher has to be a member of the NET-QUBIC consortium, or a consortium member has to collaborate in the project. The NET-QUBIC steering committee will judge the proposal. The SPSS syntax of the analyses in this study are publically available on GitHub: https://github.com/b-lissenberg/COVID-and-cancer.

## Funding

This work was supported by the Dutch Cancer Society [grant number VU 2013–5930] and the Dutch Cancer Society, Alpe Young Investigator Grant [grant number 12820]. The funding body had no role in the design of the study and collection, analysis, and interpretation of data nor in writing the manuscript.

## Competing interests

BLW: none

FJ: received funding from the Dutch Cancer Society, Alpe Young Investigator Grant [grant number 12820]. All payments were made to the institution.

RJBdJ: none

FL: received funding from the Dutch Cancer Society [grant numbers VU2017-8288 and 11839]. All payments were made to the institution.

CRL: none

SFO: received funding from Celidex Therapeutics, payment or honoraria for lectures, presentations, speakers bureaus, manuscript writing or educational events from Merck and participated in the Data Safety Monitoring Board or Advisory Board of GenMab and Bristol Myers Squibb. All payments were made to the institution.

RPT: none

IMVdL: received funding from the Dutch Cancer Society (grant number VU 2013-5930). All payments were made to the institution.

## Data availability

The dataset generated and analyzed (including a deindentified version of it) during the current study is not publicly available as the collection and integration of large amounts of personal, biological, genetic and diagnostic information precludes open access to the NET-QUBIC research data. Data are available from the corresponding author on reasonable request. On the NET-QUBIC website (www.kubusproject.nl) is described how NET-QUBIC data are made available for the research community. In short, a research proposal has to be submitted to the NET-QUBIC steering committee. The researcher has to be a member of the NET-QUBIC consortium, or a consortium member has to collaborate in the project. The NET-QUBIC steering committee will judge the proposal. The SPSS syntax of the analyses in this study are publically available on GitHub: https://github.com/b-lissenberg/COVID-and-cancer.

## Supplementary Tables

**Supplemental Table 1.** Patients characteristics at baseline unless specified otherwise for included survivors compared to excluded survivors. Data is described as number and percentage or as mean (standard deviation).

|  | included current<br>study (N=318) |  | excluded current<br>study (N=142) |  | p-value |
| --- | --- | --- | --- | --- | --- |
| Age (years) | 63.5 (9.7) |  | 63.3 (10.0) |  | 0.77 |
| Gender |  |  |  |  | 0.94 |
| Men | 234 | 73.6% | 105 | 73.9% |  |
| Women | 84 | 26.4% | 37 | 26.1% |  |
| Education level |  |  |  |  | 0.77 |
| Low | 123 | 40.3% | 58 | 43.0% |  |
| Middle | 82 | 26.9% | 32 | 23.7% |  |
| High | 100 | 32.8% | 45 | 33.3% |  |
| Missing | 13 |  | 7 |  |  |
| Living arrangement |  |  |  |  | 0.70 |
| Living together | 243 | 79.4% | 105 | 77.8% |  |
| Living alone | 63 | 20.6% | 30 | 22.2% |  |
| Missing | 12 |  | 7 |  |  |
| Marital status |  |  |  |  | 0.75 |
| not married | 107 | 35.0% | 49 | 36.6% |  |
| married | 199 | 65.0% | 85 | 63.4% |  |
| Missing | 12 |  | 8 |  |  |
| NEO-FFI extraversion<sup>a</sup> | 40.9 (6.3) |  | 39.9 (6.1) |  | 0.17 |
| Tumor location |  |  |  |  | 0.53 |
| Oral cavity | 80 | 25.2% | 40 | 28.2% |  |
| Oropharynx | 115 | 36.2% | 48 | 33.8% |  |
| Hypopharynx | 18 | 5.7% | 4 | 2.8% |  |
| Larynx | 94 | 29.6% | 47 | 33.1% |  |
| Unknown primary | 11 | 3.5% | 3 | 2.1% |  |
| Tumor stage |  |  |  |  | 0.56 |
| Stage 0(Cis)/I<sup>+</sup> | 84 | 26.4% | 38 | 26.8% |  |
| Stage II | 59 | 18.6% | 33 | 23.2% |  |
| Stage III | 50 | 15.7% | 17 | 12.0% |  |
| Stage IV | 125 | 39.3% | 54 | 38.0% |  |
| Treatment modality (dichotomized) |  |  |  |  | 0.44 |
| Single modality | 178 | 56.0% | 85 | 59.9% |  |
| Multimodality | 140 | 44.0% | 57 | 40.1% |  |
| WHO performance status |  |  |  |  | 0.20 |
| 0 | 246 | 77.4% | 102 | 71.8% |  |
| >0 | 72 | 22.6% | 40 | 28.2% |  |
| Comorbidity |  |  |  |  | 0.010 |
| None | 116 | 36.9% | 30 | 23.1% |  |
| Mild | 122 | 38.9% | 52 | 40.0% |  |
| Moderate | 54 | 17.2% | 31 | 23.8% |  |
| Severe | 22 | 7.0% | 17 | 13.1% |  |
| Missing | 4 |  | 12 |  |  |
| IADL at M24<sup>b</sup> | 7.2 (1.0) |  | 7.1 (1.2) |  | 0.25 |
| Major depressive disorder past year at M24 |  |  |  |  | 0.013 |
| No | 245 | 95.3% | 98 | 89.9% |  |
| Yes | 12 | 4.7% | 11 | 10.1% |  |
| Missing | 61 |  | 33 |  |  |
| Lifetime major depressive disorder at M24 |  |  |  |  | 0.005 |
| No | 225 | 87.5% | 66 | 65.3% |  |
| Yes | 32 | 12.5% | 35 | 34.7% |  |
| Missing | 61 |  | 41 |  |  |
| Paid work at M24 |  |  |  |  | 0.039 |
| No | 192 | 65.1% | 82 | 75.9% |  |
| Yes | 103 | 34.9% | 26 | 24.1% |  |
| Missing | 23 |  | 34 |  |  |
| Excessive alcohol consumption at M24 |  |  |  |  | 0.37 |
| No | 228 | 78.9% | 51 | 82.3% |  |
| Yes | 61 | 21.1% | 11 | 17.7% |  |
| Missing | 29 |  | 80 |  |  |
| Smoking behavior at M24 |  |  |  |  | 0.19 |
| Not a current smoker | 239 | 82.7% | 52 | 81.3% |  |
| Current smoker | 50 | 17.3% | 12 | 18.8% |  |
| Missing | 29 |  | 78 |  |  |
| BMI (kg/m2) at M24<sup>b</sup> | 26.2 (4.3) |  | 25.1 (4.3) |  | 0.074 |

\*One patient had a cTNM stage of 0, however, pTNM was stage 2.
<sup>a</sup>missing for n=32 excluded and n=15 included
<sup>b</sup>missing for n=34 excluded and n=26 included
NEO-FFI=NEO Five Factor Inventory; IADL=deze Instrumental Activities of Daily Living; BMI=body mass index

**Supplemental Table 2.** Descriptive statistics (total sample size (N), mean and standard deviation (SD)) per measurement for continuous PROMs per group.

| PROM | M36 before COVID (N=199) |  |  |  |  |  | M36 during COVID (N=119) |  |  |  |  |  |
| --- | --- | --- | --- | --- | --- | --- | --- | --- | --- | --- | --- | --- |
|  | M24 |  |  | M36 |  |  | M24 |  |  | M36 |  |  |
|  | N | mean | SD | N | mean | SD | N | mean | SD | N | mean | SD |
| **EORTC - C30 domain** |  |  |  |  |  |  |  |  |  |  |  |  |
| Global quality of life [0-100] | 189 | 79.7 | 17.0 | 192 | 81.1 | 15.8 | 118 | 79.7 | 17.0 | 108 | 79.2 | 18.8 |
| Physical functioning [0-100] | 189 | 87.7 | 14.4 | 193 | 87.7 | 16.3 | 118 | 89.3 | 14.8 | 110 | 87.8 | 17.6 |
| Role functioning [0-100] | 189 | 86.7 | 21.3 | 193 | 87.7 | 22.2 | 118 | 88.6 | 20.1 | 110 | 87.0 | 22.4 |
| Emotional functioning [0-100] | 189 | 89.0 | 16.7 | 193 | 89.6 | 15.9 | 118 | 90.8 | 14.3 | 108 | 89.1 | 15.6 |
| Cognitive functioning [0-100] | 189 | 89.7 | 15.5 | 193 | 88.9 | 16.0 | 118 | 87.6 | 16.0 | 108 | 87.8 | 15.9 |
| Social functioning [0-100] | 189 | 91.5 | 15.9 | 193 | 91.5 | 16.1 | 118 | 93.5 | 13.1 | 108 | 90.6 | 19.2 |
| Fatigue [0-100] | 189 | 20.1 | 20.6 | 192 | 21.5 | 22.6 | 118 | 21.7 | 21.3 | 110 | 21.4 | 22.0 |
| Nausea Vomiting [0-100] | 189 | 1.5 | 5.6 | 193 | 1.6 | 7.0 | 118 | 0.8 | 5.3 | 110 | 1.8 | 6.1 |
| Pain [0-100] | 189 | 13.1 | 19.1 | 193 | 11.6 | 18.5 | 118 | 12.0 | 20.2 | 110 | 12.9 | 20.9 |
| Dyspnoea [0-100] | 189 | 12.2 | 22.5 | 193 | 14.2 | 23.7 | 118 | 12.1 | 21.6 | 110 | 13.9 | 24.9 |
| Insomnia [0-100] | 189 | 16.0 | 24.7 | 192 | 17.0 | 25.7 | 118 | 20.1 | 26.6 | 110 | 19.4 | 28.0 |
| Loss of appetite [0-100] | 189 | 7.6 | 17.1 | 193 | 5.7 | 15.5 | 118 | 4.8 | 15.9 | 110 | 7.3 | 20.9 |
| Constipation [0-100] | 189 | 6.5 | 15.7 | 193 | 6.7 | 16.5 | 117 | 6.3 | 14.5 | 110 | 6.7 | 14.8 |
| Diarrhoea [0-100] | 189 | 6.0 | 15.0 | 191 | 4.2 | 13.9 | 117 | 4.3 | 12.0 | 110 | 3.3 | 10.0 |
| Financial problems [0-100] | 187 | 5.2 | 15.6 | 193 | 6.4 | 15.9 | 118 | 5.9 | 18.3 | 108 | 4.9 | 16.3 |
| EORTC summary score [0-100] | 189 | 89.4 | 9.9 | 190 | 89.7 | 10.3 | 117 | 89.9 | 10.0 | 108 | 88.8 | 12.1 |
| **EORTC - HN35 domain** |  |  |  |  |  |  |  |  |  |  |  |  |
| Pain [0-100] | 188 | 12.0 | 15.8 | 193 | 10.2 | 14.1 | 117 | 10.6 | 15.2 | 108 | 9.4 | 16.3 |
| Swallowing [0-100] | 189 | 9.7 | 14.6 | 193 | 9.3 | 13.6 | 118 | 7.8 | 14.1 | 108 | 9.1 | 16.4 |
| Senses problems [0-100] | 189 | 12.6 | 19.9 | 193 | 13.2 | 21.3 | 118 | 11.7 | 19.6 | 108 | 13.0 | 23.7 |
| Trouble with social contact [0-100] | 189 | 2.2 | 7.0 | 193 | 3.4 | 10.4 | 117 | 3.0 | 8.0 | 108 | 4.4 | 9.9 |
| Trouble with social eating [0-100] | 189 | 8.2 | 15.7 | 193 | 8.9 | 17.6 | 118 | 5.6 | 12.2 | 108 | 7.2 | 17.7 |
| Speech problems [0-100] | 189 | 9.9 | 14.7 | 193 | 10.4 | 15.5 | 118 | 9.7 | 14.6 | 108 | 8.5 | 15.5 |
| Less sexuality [0-100] | 176 | 23.7 | 32.1 | 177 | 27.6 | 34.5 | 108 | 23.8 | 31.5 | 102 | 28.4 | 32.5 |
| Teeth [0-100] | 189 | 10.8 | 21.9 | 193 | 11.7 | 22.6 | 117 | 8.5 | 18.1 | 107 | 7.5 | 17.3 |
| Opening mouth [0-100] | 189 | 8.5 | 21.7 | 193 | 10.0 | 21.6 | 118 | 9.6 | 22.7 | 108 | 8.6 | 17.9 |
| Dry mouth [0-100] | 188 | 35.5 | 32.8 | 193 | 33.2 | 30.3 | 118 | 28.5 | 28.7 | 108 | 29.3 | 26.9 |
| Sticky saliva [0-100] | 188 | 24.8 | 28.0 | 188 | 24.8 | 28.0 | 118 | 21.2 | 30.7 | 108 | 20.7 | 27.2 |
| Coughing [0-100] | 189 | 19.6 | 24.0 | 193 | 19.2 | 24.4 | 118 | 18.9 | 24.5 | 108 | 16.4 | 22.1 |
| Felt ill [0-100] | 189 | 7.4 | 17.6 | 193 | 6.6 | 15.7 | 118 | 5.4 | 13.8 | 108 | 4.0 | 11.8 |
| **PASE** |  |  |  |  |  |  |  |  |  |  |  |  |
| Leisure activity | 187 | 40.7 | 40.2 | 188 | 42.3 | 36.7 | 115 | 49.5 | 54.6 | 110 | 42.3 | 43.1 |
| Household activity | 187 | 71.6 | 59.7 | 188 | 79.6 | 79.5 | 115 | 69.9 | 64.6 | 110 | 70.4 | 62.3 |
| Work activity | 186 | 18.3 | 39.8 | 184 | 20.7 | 43.0 | 112 | 14.2 | 32.7 | 108 | 11.9 | 35.5 |
| Total activity | 187 | 130.5 | 93.5 | 188 | 142.1 | 104.6 | 115 | 133.2 | 106.4 | 110 | 124.4 | 96.1 |
| **HADS** |  |  |  |  |  |  |  |  |  |  |  |  |
| Depression [0-21] | 187 | 2.5 | 2.9 | 188 | 2.5 | 3.2 | 118 | 2.6 | 3.2 | 110 | 2.9 | 3.3 |
| Anxiety [0-21] | 188 | 3.0 | 3.1 | 189 | 3.0 | 3.2 | 118 | 3.0 | 3.1 | 108 | 2.8 | 2.9 |
| Total score [0-42] | 186 | 5.5 | 5.4 | 187 | 5.5 | 5.7 | 118 | 5.6 | 5.9 | 108 | 5.6 | 5.4 |
| **CWS** |  |  |  |  |  |  |  |  |  |  |  |  |
| Fear of recurrence [8-32] | 184 | 11.8 | 4.0 | 187 | 11.6 | 3.5 | 114 | 11.5 | 3.4 | 108 | 11.4 | 4.2 |
| **De Jong Gierveld** |  |  |  |  |  |  |  |  |  |  |  |  |
| Emotional loneliness score [0-6] | 180 | 1.0 | 1.6 | 185 | 1.1 | 1.8 | 112 | 1.1 | 1.8 | 109 | 1.3 | 1.9 |
| Social loneliness score [0-5] | 184 | 1.4 | 1.7 | 185 | 1.6 | 1.8 | 113 | 1.5 | 1.8 | 107 | 1.5 | 1.7 |
| Total loneliness score [0-11] | 184 | 2.4 | 3.0 | 186 | 2.7 | 3.1 | 114 | 2.6 | 3.2 | 109 | 2.7 | 3.1 |

**Supplemental Table 3.** Descriptive statistics (frequency (n) and percentage) per assessment for dichotomous and categorical PROMs per group

|  |  |  | M36 before COVID (N=199) |  |  |  | M36 during COVID (N=119) |  |  |  |
| --- | --- | --- | --- | --- | --- | --- | --- | --- | --- | --- |
|  |  |  | M24 |  | M36 |  | M24 |  | M36 |  |
|  |  |  | n | % | n | % | n | % | n | % |
| **EORTC - HN35 domain** |  |  |  |  |  |  |  |  |  |  |
| Painkillers | no |  | 135 | 71.8% | 140 | 72.9% | 84 | 71.8% | 81 | 73.6% |
|  | yes |  | 53 | 28.2% | 52 | 27.1% | 33 | 28.2% | 29 | 26.4% |
| Nutritional supplements | no |  | 170 | 90.4% | 173 | 89.6% | 106 | 91.4% | 100 | 90.9% |
|  | yes |  | 18 | 9.6% | 20 | 10.4% | 10 | 8.6% | 10 | 9.1% |
| Feeding tube | no |  | 188 | 99.5% | 191 | 99.0% | 116 | 100.0% | 109 | 99.1% |
|  | yes |  | 1 | 0.5% | 2 | 1.0% | 0 | 0.0% | 1 | 0.9% |
| Weight loss | no |  | 178 | 94.7% | 179 | 92.7% | 113 | 96.6% | 99 | 90.0% |
|  | yes |  | 10 | 5.3% | 14 | 7.3% | 4 | 3.4% | 11 | 10.0% |
| Weight gain | no |  | 148 | 79.6% | 157 | 82.2% | 94 | 81.7% | 95 | 88.8% |
|  | yes |  | 38 | 20.4% | 34 | 17.8% | 21 | 18.3% | 12 | 11.2% |
| **PASE** |  |  |  |  |  |  |  |  |  |  |
| Total activity group | very poor |  | 26 | 13.9% | 33 | 17.6% | 25 | 21.7% | 30 | 27.3% |
|  | poor |  | 59 | 31.6% | 43 | 22.9% | 27 | 23.5% | 24 | 21.8% |
|  | fair |  | 41 | 21.9% | 39 | 20.7% | 25 | 21.7% | 17 | 15.5% |
|  | good |  | 24 | 12.8% | 34 | 18.1% | 16 | 13.9% | 17 | 15.5% |
|  | very good |  | 18 | 9.6% | 15 | 8.0% | 8 | 7.0% | 12 | 10.9% |
|  | excelent |  | 19 | 10.2% | 24 | 12.8% | 14 | 12.2% | 10 | 9.1% |
| **De Jong Gierveld** |  |  |  |  |  |  |  |  |  |  |
| Total loneliness category | not lonely [0-2] |  | 117 | 63.6% | 117 | 62.9% | 74 | 64.9% | 65 | 59.6% |
|  | moderate [3-8] |  | 55 | 29.9% | 52 | 28.0% | 33 | 28.9% | 35 | 32.1% |
|  | severe [9-10] |  | 10 | 5.4% | 15 | 8.1% | 2 | 1.8% | 8 | 7.3% |
|  | very severe[11] |  | 2 | 1.1% | 2 | 1.1% | 5 | 4.4% | 1 | 0.9% |

**Supplemental Table 4.** P-values for the three-way interaction between time, group and each potential influencing factor, to identify potential modifiers of the effect on the change of PROMs between 24 months and 36 months follow-up. Factors as assessed at baseline unless specified otherwise

| PROM | Age | Gender | Educational level | Living arrangement | Marital status (T0) | NEO-FFI extraversion |
| --- | --- | --- | --- | --- | --- | --- |
| **EORTC - C30 domain** |  |  |  |  |  |  |
| Global quality of life [0-100] | y0.44 | 0.34 | 0.71 | 0.49 | 0.34 | 0.62 |
| Physical functioning [0-100] | 0.52 | 0.015 | 0.59 | 0.30 | 0.05 | 0.24 |
| Role functioning [0-100] | 0.45 | 0.24 | 0.88 | 0.88 | 0.44 | 0.11 |
| Emotional functioning [0-100] | 0.74 | 0.13 | 0.44 | 0.52 | 0.85 | 0.27 |
| Cognitive functioning [0-100] | 0.61 | 0.69 | 0.73 | 0.94 | 0.96 | 0.38 |
| Social functioning [0-100] | 0.023 | 0.22 | 0.29 | 0.54 | 0.70 | 0.99 |
| Fatigue [0-100] | 0.35 | 0.84 | 0.45 | 0.99 | 0.94 | 0.091 |
| Nausea Vomiting [0-100] | 0.94 | 0.63 | 0.22 | 0.73 | 0.82 | 0.75 |
| Pain [0-100] | 0.70 | 0.45 | 0.74 | 0.69 | 0.35 | 0.96 |
| Dyspnoea [0-100] | 0.58 | 0.34 | 0.92 | 0.69 | 0.29 | 0.46 |
| Insomnia [0-100] | 0.63 | 0.43 | 0.85 | 0.77 | 0.89 | 0.71 |
| Loss of appetite [0-100] | 0.082 | 0.60 | 0.25 | 0.12 | 0.92 | 0.53 |
| Constipation [0-100] | 0.16 | 0.27 | 0.59 | 0.028 | 0.11 | 0.31 |
| Diarrhoea [0-100] | 0.11 | 0.58 | 0.37 | 0.84 | 0.39 | 0.88 |
| Financial problems [0-100] | 0.38 | 0.79 | 0.23 | 0.60 | 0.41 | 0.31 |
| EORTC summary score [0-100] | 0.59 | 0.089 | 0.72 | 0.72 | 0.63 | 0.16 |
| **EORTC - HN35 domain** |  |  |  |  |  |  |
| Pain [0-100] | 0.75 | 0.59 | 0.53 | 0.18 | 0.78 | 0.80 |
| Swallowing [0-100] | 0.83 | 0.97 | 0.83 | 0.55 | 0.42 | 0.098 |
| Senses problems [0-100] | 0.58 | 0.11 | 0.91 | 0.093 | 0.05 | 0.73 |
| Trouble with social contact [0-100] | 0.67 | 0.29 | 0.11 | 0.020 | 0.037 | 0.25 |
| Trouble with social eating [0-100] | 0.91 | 0.77 | 0.94 | 0.51 | 0.14 | 0.21 |
| Speech problems [0-100] | 0.81 | 0.13 | 0.59 | 0.38 | 0.79 | 0.70 |
| Less sexuality [0-100] | 0.48 | 0.036 | 0.13 | 0.39 | 0.18 | 0.92 |
| Teeth [0-100] | 0.51 | 0.17 | 0.51 | 0.97 | 0.61 | 0.87 |
| Opening mouth [0-100] | 0.77 | 0.98 | 0.59 | 0.45 | 0.64 | 0.74 |
| Dry mouth [0-100] | 0.58 | 0.54 | 0.65 | 0.75 | 0.72 | 0.20 |
| Sticky saliva [0-100] | 0.84 | 0.90 | 0.29 | 0.97 | 0.45 | 0.57 |
| Coughing [0-100] | 0.051 | 0.77 | 0.61 | 0.005 | 0.040 | 0.23 |
| Felt ill [0-100] | 0.22 | 0.45 | 0.92 | 0.83 | 0.61 | 0.07 |
| Painkillers | 0.53 | 0.62 | 0.67 | 0.004 | 0.015 | 0.92 |
| Nutritional supplements | 0.45 | 0.15 | 0.30 | 0.37 | 0.63 | 0.44 |
| Feeding tube | n.e. | n.e. | n.e. | n.e. | n.e. | n.e. |
| Weight loss | 0.67 | n.e. | <0.001 | n.e. | 0.77 | 1.00 |
| Weight gain | 0.17 | 0.91 | 0.34 | 0.33 | 0.82 | 0.19 |
| **PASE** |  |  |  |  |  |  |
| Leisure activity | 0.48 | 0.13 | 0.54 | 0.37 | 0.19 | 0.24 |
| Household activity | 0.87 | 0.08 | 0.21 | 0.56 | 0.36 | 0.16 |
| Work activity | 0.60 | 0.43 | 0.39 | 0.68 | 0.37 | 0.80 |
| Total activity | 0.85 | 0.033 | 0.20 | 0.36 | 0.14 | 0.70 |
| Total activity (categorical) | 0.15 | 0.018 | 0.34 | 0.55 | 0.11 | 0.99 |
| **HADS** |  |  |  |  |  |  |
| Depression [0-21] | 0.022 | 0.094 | 0.58 | 0.35 | 0.15 | 0.80 |
| Anxiety [0-21] | 0.10 | 0.021 | 0.30 | 0.42 | 0.19 | 0.27 |
| Total score [0-42] | 0.023 | 0.070 | 0.43 | 0.34 | 0.12 | 0.43 |
| **CWS** |  |  |  |  |  |  |
| Fear of recurrence [8-32] | 0.09 | 0.67 | 0.045 | 0.96 | 0.88 | 0.85 |
| **De Jong Gierveld** |  |  |  |  |  |  |
| Emotional loneliness score [0-6] | 0.82 | 0.92 | 0.33 | 0.72 | 0.49 | 0.61 |
| Social loneliness score [0-5] | 0.26 | 0.61 | 0.57 | 0.56 | 0.09 | 0.78 |
| Total loneliness score [0-11] | 0.24 | 0.88 | 0.25 | 0.96 | 0.67 | 0.70 |
| Total loneliness score (categorical) | 0.18 | 0.82 | 0.16 | 0.96 | 0.36 | 0.89 |

Supplemental Table 4 (continued).
| PROM | Tumor location | Tumor stage | Treatment modality | Comorbidity | WHO performance status | Cancer progression status (M24) |
| --- | --- | --- | --- | --- | --- | --- |
| **EORTC - C30 domain** |  |  |  |  |  |  |
| Global quality of life [0-100] | 0.15 | 0.43 | 0.84 | 0.84 | 0.73 | 0.41 |
| Physical functioning [0-100] | 0.31 | 0.96 | 0.88 | 0.19 | 0.56 | 0.78 |
| Role functioning [0-100] | 0.54 | 0.015 | 0.51 | 0.09 | 0.36 | 0.20 |
| Emotional functioning [0-100] | 0.66 | 0.21 | 0.14 | 0.73 | 0.57 | 0.028 |
| Cognitive functioning [0-100] | 0.051 | 0.87 | 0.31 | 0.030 | 0.87 | 0.35 |
| Social functioning [0-100] | 0.27 | 0.82 | 0.62 | 0.095 | 0.73 | 0.22 |
| Fatigue [0-100] | 0.69 | 0.42 | 0.26 | 0.58 | 0.56 | 0.24 |
| Nausea Vomiting [0-100] | 0.23 | 0.99 | 0.89 | 0.39 | 0.18 | 0.43 |
| Pain [0-100] | 0.89 | 0.94 | 0.88 | 0.14 | 0.59 | 0.78 |
| Dyspnoea [0-100] | 0.76 | 0.57 | 0.73 | 0.22 | 0.80 | 0.09 |
| Insomnia [0-100] | 0.61 | 0.28 | 0.96 | 0.70 | 0.94 | 0.15 |
| Loss of appetite [0-100] | 0.048 | 0.60 | 0.046 | 0.003 | 0.75 | 0.63 |
| Constipation [0-100] | 0.86 | 0.07 | 0.26 | 0.19 | 0.79 | 0.22 |
| Diarrhoea [0-100] | 0.098 | 0.09 | 0.077 | 0.74 | 0.26 | 1.00 |
| Financial problems [0-100] | 0.69 | 0.40 | 0.60 | 0.21 | 0.004 | 0.001 |
| EORTC summary score [0-100] | 0.21 | 0.13 | 0.38 | 0.18 | 0.90 | 0.95 |
| **EORTC - HN35 domain** |  |  |  |  |  |  |
| Pain [0-100] | 0.017 | 0.32 | 0.78 | 0.23 | 0.87 | 0.015 |
| Swallowing [0-100] | 0.69 | 0.96 | 0.90 | <0.001 | 0.020 | 0.010 |
| Senses problems [0-100] | 0.92 | 0.07 | 0.13 | 0.066 | 0.089 | 0.034 |
| Trouble with social contact [0-100] | 0.22 | 0.74 | 0.29 | 0.33 | 0.52 | 0.29 |
| Trouble with social eating [0-100] | 0.94 | 0.74 | 0.62 | 0.039 | 0.89 | 0.14 |
| Speech problems [0-100] | 0.15 | 0.42 | 0.20 | 0.95 | 0.92 | 0.93 |
| Less sexuality [0-100] | 0.88 | 0.63 | 0.47 | 0.026 | 0.48 | 0.31 |
| Teeth [0-100] | 0.53 | 1.00 | 0.85 | 0.083 | 0.22 | 0.18 |
| Opening mouth [0-100] | 0.091 | 0.30 | 0.90 | 0.76 | 0.38 | 0.74 |
| Dry mouth [0-100] | 0.007 | 0.78 | 0.69 | 0.37 | 0.52 | 0.25 |
| Sticky saliva [0-100] | 0.48 | 0.56 | 0.75 | 0.35 | 0.82 | 0.035 |
| Coughing [0-100] | 0.12 | 0.17 | 0.34 | 0.085 | 0.64 | 0.56 |
| Felt ill [0-100] | 0.42 | 0.30 | 0.58 | 0.15 | 0.13 | 0.63 |
| Painkillers | 0.14 | 0.40 | 0.17 | 0.13 | 0.14 | 0.32 |
| Nutritional supplements | n.e. | 0.42 | 0.31 | <0.001 | 0.55 | 0.66 |
| Feeding tube | n.e. | n.e. | n.e. | n.e. | n.e. | n.e. |
| Weight loss | 0.77 | <0.001 | 0.23 | <0.001 | 0.69 | 0.60 |
| Weight gain | 1.00 | 0.78 | 0.73 | 0.57 | 0.21 | 0.35 |
| **PASE** |  |  |  |  |  |  |
| Leisure activity | 0.63 | 0.007 | 0.36 | 0.98 | 0.46 | 0.82 |
| Household activity | 0.07 | 0.97 | 0.49 | 0.94 | 0.06 | 0.44 |
| Work activity | 1.00 | 0.83 | 0.44 | 0.61 | 0.58 | 0.50 |
| Total activity | 0.22 | 0.33 | 0.88 | 0.94 | 0.19 | 0.85 |
| Total activity (categorical) | 0.003 | 0.48 | 0.81 | 0.27 | 0.07 | 0.90 |
| **HADS** |  |  |  |  |  |  |
| Depression [0-21] | 0.77 | 0.76 | 0.022 | 0.35 | 0.74 | 0.64 |
| Anxiety [0-21] | 0.53 | 0.55 | 0.51 | 0.46 | 0.99 | 0.42 |
| Total score [0-42] | 0.64 | 0.99 | 0.035 | 0.55 | 0.77 | 0.39 |
| **CWS** |  |  |  |  |  |  |
| Fear of recurrence [8-32] | 0.65 | 0.051 | 0.30 | 0.51 | 0.14 | 0.64 |
| **De Jong Gierveld** |  |  |  |  |  |  |
| Emotional loneliness score [0-6] | 0.48 | 0.24 | 0.023 | 0.54 | 0.83 | 0.045 |
| Social loneliness score [0-5] | 0.36 | 0.25 | 0.65 | 0.79 | 0.22 | 0.63 |
| Total loneliness score [0-11] | 0.43 | 0.08 | 0.09 | 0.50 | 0.44 | 0.10 |
| Total loneliness score (categorical) | 0.30 | 0.64 | 0.65 | 0.39 | 0.92 | 0.20 |

Supplemental Table 4 (continued).
| PROM | IADL (M24) | Major depressive disorder past year (M24) | Lifetime major depressive disorder (M24) | Paid work (M24) | Excessive alcohol consumption (M24) | Smoking behavior (M24) | BMI (M24) |
| --- | --- | --- | --- | --- | --- | --- | --- |
| **EORTC - C30 domain** |  |  |  |  |  |  |  |
| Global quality of life [0-100] | 0.93 | 0.57 | 0.30 | 0.49 | 0.61 | 0.21 | 0.66 |
| Physical functioning [0-100] | 0.56 | 0.29 | 0.20 | 0.24 | 0.66 | 0.71 | 0.37 |
| Role functioning [0-100] | 1.00 | 0.014 | 0.24 | 0.42 | 0.47 | 0.015 | 0.30 |
| Emotional functioning [0-100] | 0.069 | 0.61 | 0.63 | 0.80 | 0.72 | 0.090 | 0.56 |
| Cognitive functioning [0-100] | 0.84 | 0.28 | 0.40 | 0.46 | 0.71 | 0.11 | 0.12 |
| Social functioning [0-100] | 0.88 | 0.10 | 0.55 | 0.19 | 0.69 | 0.47 | 0.92 |
| Fatigue [0-100] | 0.053 | 0.73 | 0.91 | 0.09 | 0.35 | 0.41 | 0.20 |
| Nausea Vomiting [0-100] | 0.95 | 0.40 | 0.37 | 0.15 | 0.26 | 0.13 | 0.74 |
| Pain [0-100] | 0.66 | 0.92 | 0.81 | 0.74 | 0.51 | 0.91 | 0.44 |
| Dyspnoea [0-100] | 0.24 | 0.95 | 0.84 | 0.79 | 0.60 | 0.15 | 0.20 |
| Insomnia [0-100] | 0.065 | 0.82 | 0.80 | 0.66 | 0.20 | 0.46 | 0.36 |
| Loss of appetite [0-100] | 0.68 | 0.99 | 0.30 | 0.35 | 0.34 | 0.79 | 0.19 |
| Constipation [0-100] | 0.88 | 0.35 | 0.32 | 0.71 | 0.28 | 0.62 | 0.26 |
| Diarrhoea [0-100] | 0.078 | 0.89 | 0.10 | 0.020 | 0.69 | 0.16 | 0.53 |
| Financial problems [0-100] | 0.008 | 0.64 | 0.84 | 0.21 | 0.64 | 0.69 | 0.23 |
| EORTC summary score [0-100] | 0.97 | 0.96 | 0.11 | 0.60 | 0.44 | 0.99 | 0.81 |
| **EORTC - HN35 domain** |  |  |  |  |  |  |  |
| Pain [0-100] | 0.90 | 0.16 | 0.81 | 0.57 | 0.73 | 0.68 | 0.020 |
| Swallowing [0-100] | 0.32 | 0.63 | 0.11 | 0.72 | 0.74 | 0.60 | 0.58 |
| Senses problems [0-100] | 0.37 | 0.78 | 0.78 | 0.47 | 0.13 | 0.89 | 0.30 |
| Trouble with social contact [0-100] | 0.99 | 0.001 | 0.027 | 0.056 | 1.00 | 0.003 | 0.25 |
| Trouble with social eating [0-100] | 0.39 | 0.20 | 0.42 | 0.41 | 0.23 | 0.30 | 0.18 |
| Speech problems [0-100] | 0.90 | 0.78 | 0.90 | 0.64 | 0.43 | 0.86 | 0.39 |
| Less sexuality [0-100] | 0.36 | 0.19 | 0.023 | 0.76 | 0.30 | 0.83 | 0.16 |
| Teeth [0-100] | 0.63 | 0.84 | 0.84 | 0.34 | 0.97 | 0.52 | 0.19 |
| Opening mouth [0-100] | 0.65 | 0.29 | 0.011 | 0.36 | 0.49 | 0.99 | 0.072 |
| Dry mouth [0-100] | 0.058 | 0.51 | 0.25 | 0.51 | 0.92 | 0.78 | 0.72 |
| Sticky saliva [0-100] | 0.53 | 0.34 | 0.50 | 0.39 | 0.59 | 0.046 | 0.096 |
| Coughing [0-100] | 0.074 | 0.54 | 0.11 | 0.14 | 0.64 | 0.016 | 0.16 |
| Felt ill [0-100] | 0.66 | 0.09 | 0.38 | 0.42 | 0.31 | 0.37 | 0.64 |
| Painkillers | 0.14 | 0.28 | 0.79 | 0.92 | 0.28 | 0.14 | 0.099 |
| Nutritional supplements | 0.20 | n.e. | n.e. | 0.61 | 0.53 | 0.64 | 0.10 |
| Feeding tube | n.e. | n.e. | n.e. | n.e. | n.e. | n.e. | n.e. |
| Weight loss | 0.77 | n.e. | <0.001 | 1.00 | n.e. | n.e. | 0.32 |
| Weight gain | 0.38 | 0.47 | 0.32 | 0.16 | 0.35 | 0.044 | 0.40 |
| **PASE** |  |  |  |  |  |  |  |
| Leisure activity | 0.50 | 0.96 | 0.034 | 0.64 | 0.18 | 0.43 | 0.81 |
| Household activity | 0.12 | 0.34 | 0.65 | 0.30 | 0.33 | 0.81 | 0.60 |
| Work activity | 0.34 | 0.53 | 1.00 | 0.22 | 0.86 | 0.38 | 0.058 |
| Total activity | 0.30 | 0.70 | 0.20 | 0.19 | 0.25 | 0.85 | 0.22 |
| Total activity (categorical) | 0.27 | 1.00 | 0.18 | 0.31 | 0.50 | 0.50 | 0.24 |
| **HADS** |  |  |  |  |  |  |  |
| Depression [0-21] | 0.53 | 0.39 | 0.35 | 0.33 | 0.21 | 0.68 | 0.33 |
| Anxiety [0-21] | 0.65 | 0.39 | 0.16 | 0.24 | 0.40 | 0.16 | 0.20 |
| Total score [0-42] | 0.87 | 0.27 | 0.18 | 0.14 | 0.83 | 0.35 | 0.18 |
| **CWS** |  |  |  |  |  |  |  |
| Fear of recurrence [8-32] | 0.22 | 0.07 | 0.007 | 0.73 | 0.57 | 0.93 | 0.061 |
| **De Jong Gierveld** |  |  |  |  |  |  |  |
| Emotional loneliness score [0-6] | 0.29 | 0.07 | 0.004 | 0.63 | 0.43 | 0.59 | 0.003 |
| Social loneliness score [0-5] | 0.95 | 0.60 | 0.85 | 0.44 | 0.50 | 0.96 | 0.32 |
| Total loneliness score [0-11] | 0.46 | 0.58 | 0.09 | 0.38 | 0.33 | 0.65 | 0.010 |
| Total loneliness score (categorical) | 0.29 | 1.00 | 0.90 | 0.27 | 0.53 | 0.96 | 0.21 |

n.e.=not estimable due to low number of events

**Supplemental Table 5A.**
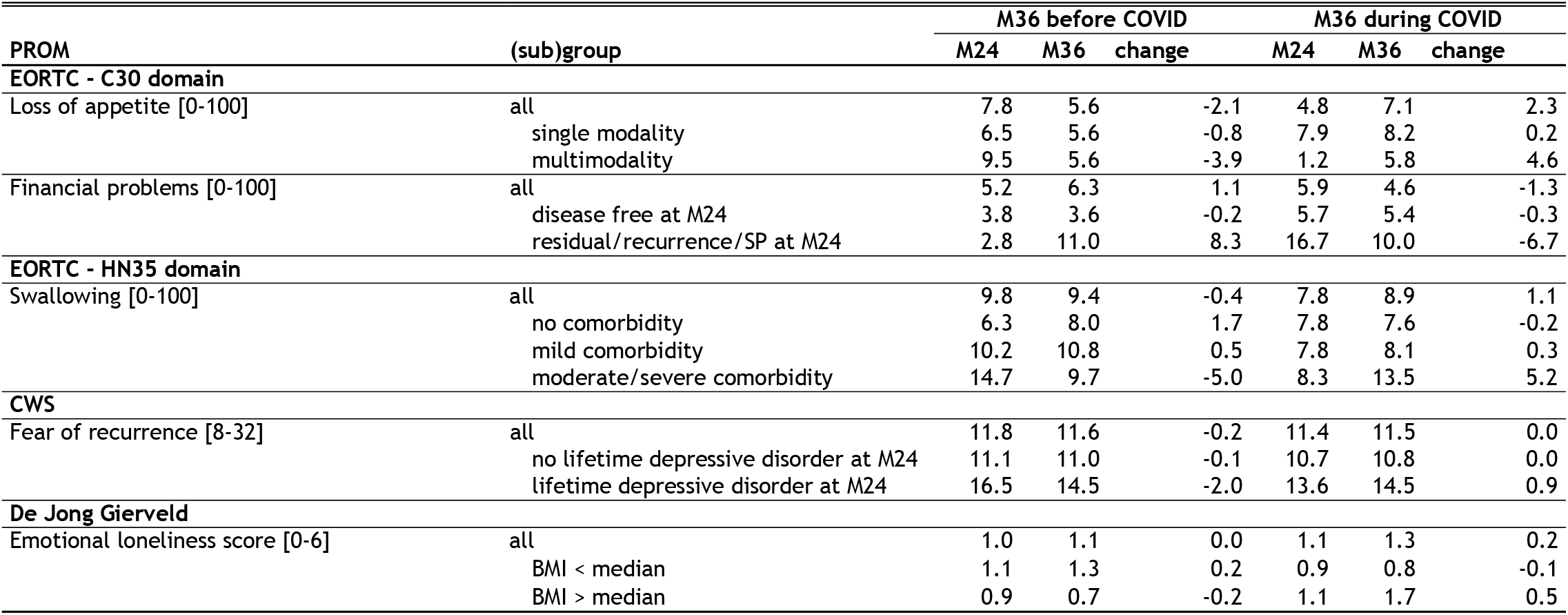
Stratified estimated marginal means for continous PROMs per assessment and per group, with corresponding estimated changes between 24 (M24) and 36 (M36) months follow-up.

**Supplemental Table 5B.**
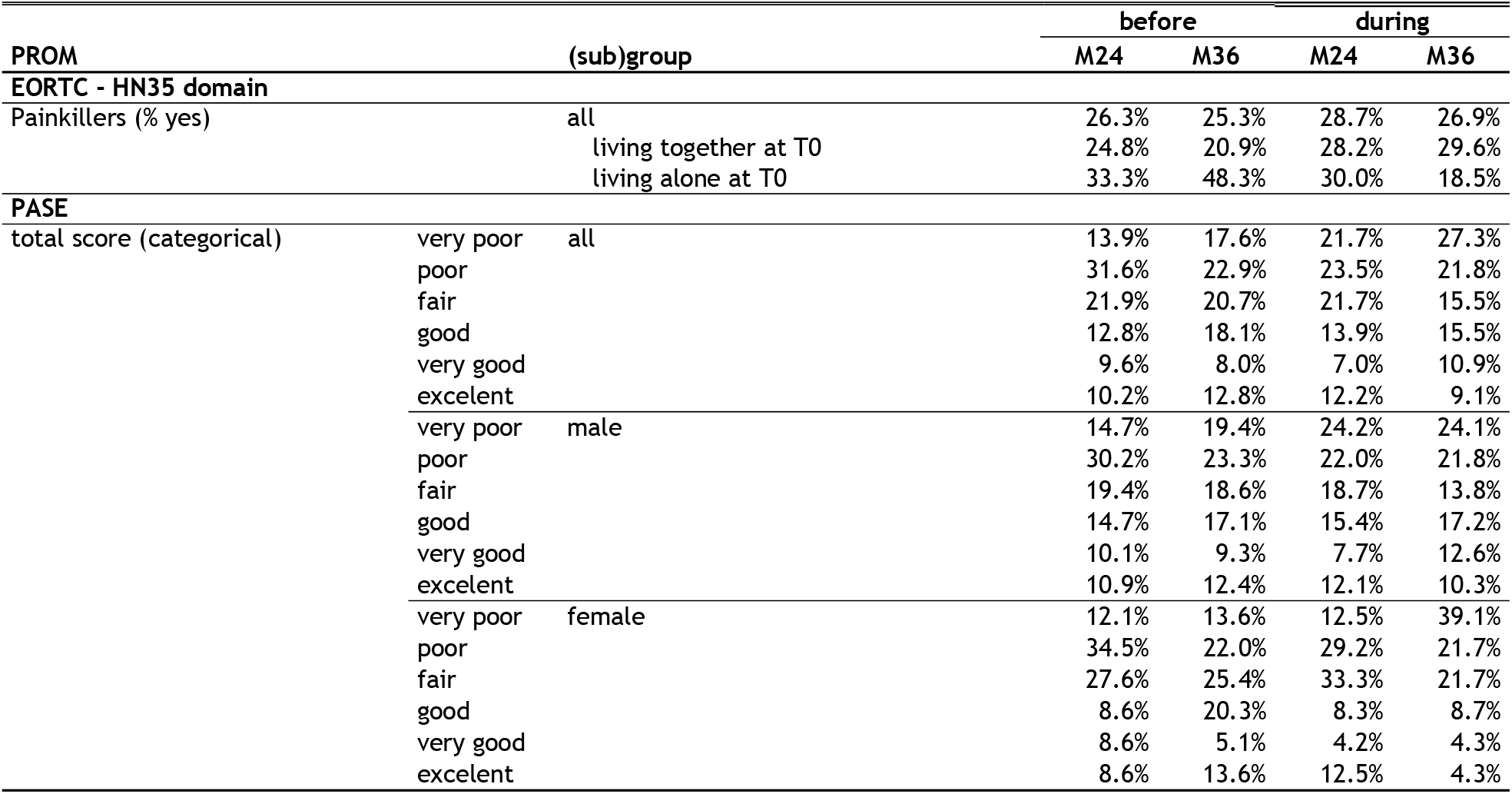
Stratified observed percentage for dichotomous and categorical PROMs per assessment and per group.

## Notes

### Competing Interest Statement

FJ received funding from the Dutch Cancer Society, Alpe Young Investigator Grant [grant number 12820]. FL received funding from the Dutch Cancer Society [grant numbers VU2017-8288 and 11839]. SFO received funding from Celidex Therapeutics, payment or honoraria for lectures, presentations, speakers bureaus, manuscript writing or educational events from Merck and participated in the Data Safety Monitoring Board or Advisory Board of GenMab and Bristol Myers Squibb. IMVdL received funding from the Dutch Cancer Society (grant number VU 2013-5930). All payments were made to the respective institutions. All other authors report no competing interests.

### Author Declarations

Medical Ethical Committee of Amsterdam UMC location VUmc (METc VUmc 2013.301 (A2018.307)-NL45051.029.13)

